# Homebound by COVID19: The Benefits and Consequences of Non-Pharmaceutical Intervention Strategies

**DOI:** 10.1101/2020.07.22.20160085

**Authors:** Buse Eylul Oruc, Arden Baxter, Pinar Keskinocak, John Asplund, Nicoleta Serban

## Abstract

**Objectives:** To evaluate the tradeoffs between potential benefits (e.g., reduction in infection spread and deaths) of non-pharmaceutical interventions for COVID19 and being homebound (i.e., refraining from community/workplace interactions).

**Methods:** An agent-based simulation model to project the disease spread and estimate the number of homebound people and person-days under multiple scenarios, including combinations of shelter-in- place, voluntary quarantine, and school closure in Georgia from March 1 to September 1, 2020.

**Results:** Compared to no intervention, under voluntary quarantine, voluntary quarantine with school closure, and shelter-in-place with school closure scenarios 3.43, 19.8, and 200+ homebound adult-days were required to prevent one infection, with the maximum number of adults homebound on a given day in the range of 121K-268K, 522K-567K, 5,377K-5,380K, respectively.

**Conclusions:** Voluntary quarantine combined with school closure significantly reduced the number of infections and deaths with a considerably smaller number of homebound person-days compared to shelter-in-place.

**Three-question Summary Box:**

1. What is the current understanding of this subject? Recent research has been conducted by various countries and regions on the impact of non-pharmaceutical interventions (NPIs) on reducing the spread of COVID19.
2. What does this report add to the literature? Our report assessed which intervention strategies provided the best results in terms of both reducing infection outcomes (cases, deaths, etc.) and minimizing their social and economic effects (e.g., number of people homebound, providing childcare, etc.).
3. What are the implications for public health practice? Voluntary quarantine proved to be the most beneficial in terms of reducing infections and deaths compared to the number of people who were homebound.

## INTRODUCTION

Recent research and experiences from various communities around the world highlighted the potential benefits of non-pharmaceutical interventions (NPIs) for slowing down the spread of COVID19 and reducing the severe health outcomes ^1-3^. NPIs include school closure, reducing public gatherings, social distancing, restricting travel, and voluntary quarantine (entire household staying at home if someone in the household has symptoms) ^4-7^ and more stringent interventions such as shelter-in-place ^8,9^.

People may become “homebound” (i.e., stay home and refrain from interactions in the community/workplace) due to complying with some of the NPIs (even if they do not experience symptoms), showing symptoms, or providing childcare. Hence, despite their benefits, there are also unintended consequences of NPIs, including the impact on the economy, unemployment, household spending, mobility, energy usage, etc. ^10-13^ and the social impact on caring for the elderly, education of the young, family support, domestic violence, and personal health and wellbeing ^14-23^.

Some NPIs, such as shelter-in-place, apply to large populations for an extended duration, whereas others, such as voluntary quarantine, impact targeted populations for a limited time. It is important to understand the tradeoffs between the public health benefits and other consequences of NPIs, particularly, as measured by homebound person-days or the size of the homebound population over time. There is sparse research on assessing which interventions have a higher overall impact in reducing societal interactions versus the ability to reduce infection spread and adverse outcomes ^8,9,24,25^.

This study evaluates the trade-offs between the public health impact measures (e.g., the number of cases, hospitalizations and deaths ^26^) and intervention metrics, including number of homebound people and person-days under various NPI scenarios, including variations of shelter-in-place, voluntary quarantine, and school closure. The intervention metrics aim to capture how much an intervention reduces societal activity and interaction, much needed to maintain economic and social life. Such evaluations can assist local and national decision makers in choosing different combinations of targeted interventions over time to reduce infection spread while considering the societal and economic impact.

## METHODS

### Intervention Analysis

The following NPIs, with varying combinations and compliance levels in different scenarios (*Figure 1*), are analyzed in this study and compared to the baseline of no intervention (NI):

**Figure 1:**
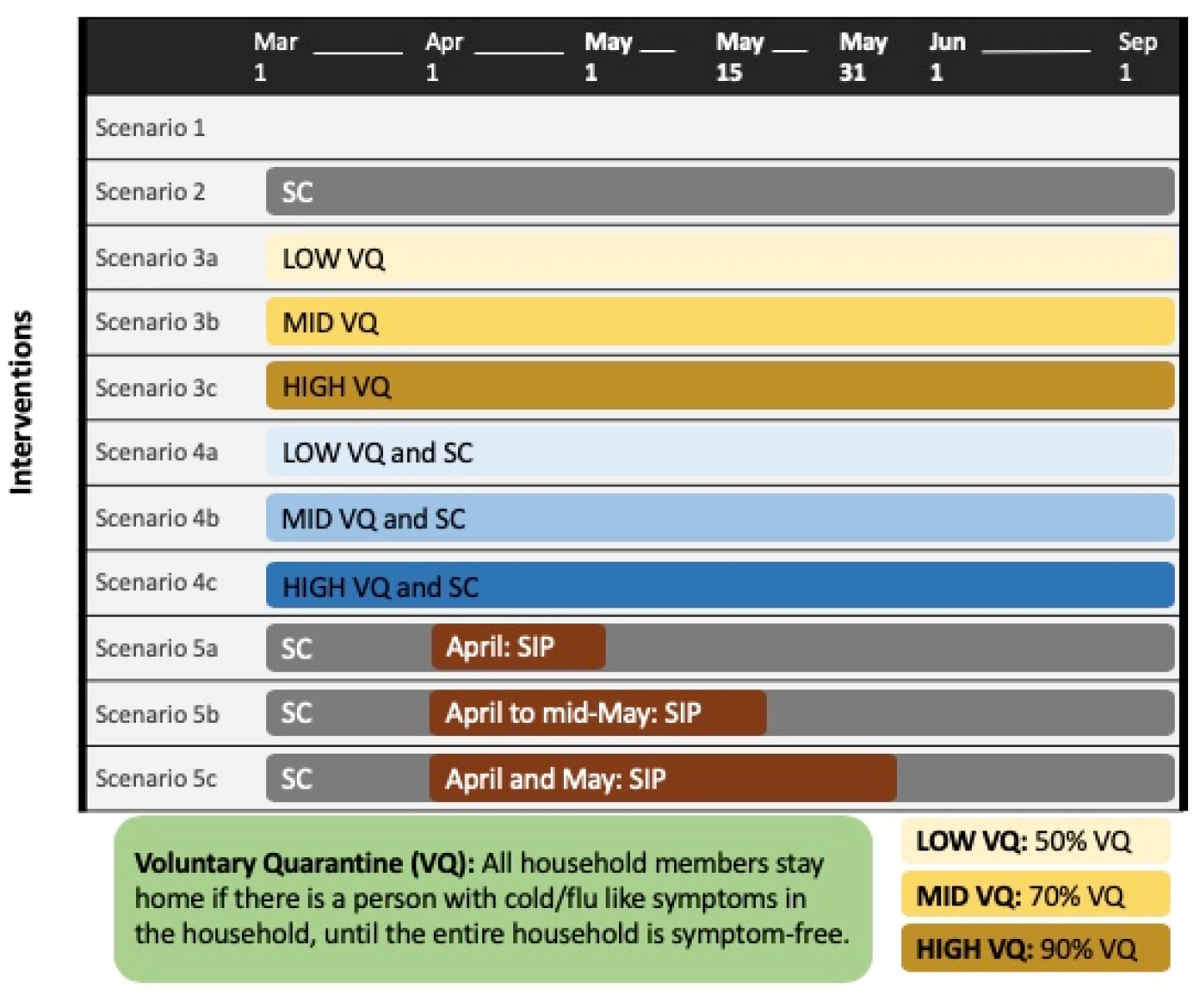
Description of the intervention scenarios considered in this study.

1. *<u>School Closure (SC)</u>* – No peer-group interactions among children or youth (i.e., no K-12 school interactions).
2. *<u>Voluntary Quarantine (VQ)</u>* –Household members stay home if any member of the household is symptomatic, until the entire household is symptom-free.
3. *<u>Shelter-in-Place (SIP)</u>* – Household members stay home.

### Modeling Case Projection and Estimating Intervention Impact

An agent-based simulation model with heterogeneous population mixing was utilized and adapted, which has been previously applied to project the number of COVID19 infections and severe outcomes under various social distancing strategies ^26^. The study period is March 1, 2020-September 1, 2020.

The population in the simulation includes children (ages 0-9), youth (ages 10-19), adults (ages 20-64), and elderly (ages 65+). The simulation monitors the health status (e.g., symptomatic, hospitalized, dead) as well as the homebound status of each household member (further details are provided in *Supplementary Material Section A* and *Table B1*).

- <u>Homebound:</u> For adults and elderly, this status is defined as staying home due to voluntary quarantine, symptoms, shelter-in-place, or *at home childcare*, i.e., providing supervision to a child who is home due to their status (e.g., due to symptoms or school closure). For example, if a child is at home in need of supervision, the status of an adult or elderly member in the household is updated to indicate that they provide supervision, labeled as *at home childcare*. For children and youth, homebound is defined as staying home due to voluntary quarantine, symptoms, or school closure.
- <u>Inactive:</u> For adults and elderly, a status of inactive refers to being inactive from society due to being homebound, hospitalized, or providing hospital care, i.e., caring for a child or youth who became hospitalized. A status of inactive for children and youth is defined as being inactive from society due to being homebound or hospitalized.

### Infection Spread Outcome Measures and Intervention Metrics

The infection spread outcome measures reported for the study period include:

- <u>Cumulative deaths</u>: Number of people who died due to COVID19.
- <u>Cumulative infections</u>: Number of people infected (including asymptomatic infections).
- <u>Peak day</u>: The day when the number of new infections was highest.
- <u>Peak infection</u>: The highest number (or percentage) of the population infected on a given day.

A statistical summary of infection spread outcome measures under baseline and intervention scenarios is provided in *Supplementary Material Table B2*.

The infection spread measures are contrasted with the following intervention metrics, which are reported for the study period:

- <u>Homebound or inactive subpopulation</u>: Number of people in a subpopulation (adults/elderly or children/youth) with homebound or inactive status, respectively, on a given day.
- <u>Percentage of days adults homebound or inactive</u>: Average percentage of days an adult has homebound or inactive status, respectively.
- <u>Homebound days:</u> Average number of days a (sub)population has homebound status.
- <u>Homebound or inactive peak day:</u> The day when the number of a (sub)population has homebound or inactive status, respectively, is highest.
- <u>Homebound or inactive peak:</u> The highest number (or percentage) of a (sub)population homebound or inactive, respectively, on a given day.
- <u>Adults absent from work:</u> The number of adults who are absent from work due to an inactive status (further details are provided in *Supplementary Material Section B*).
- <u>Homebound days to prevent an infection</u>: Additional adult homebound days needed to prevent an infection (in Scenario X, relative to Scenario 1), calculated as follows:

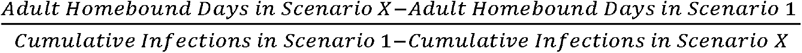
- <u>Homebound days to prevent a death</u>: Additional adult homebound days needed to prevent a death (in Scenario X, relative to Scenario 1), calculated as follows:

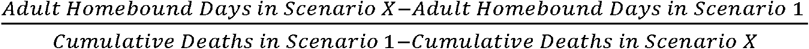

## RESULTS

*Supplementary Material Table B2* presents the infection spread outcome measures, including the population infected or dead and the peak infection.

*Figure 2* presents the daily new infections and the homebound adults over time across all scenarios. Under Scenarios 1, 3a, 3b, 3c (non-school closure scenarios), the homebound peak for adults decreased from 267,566 under Scenario 3a to 121,346 under Scenario 3c, and the peak under Scenario 1 was 242,948. Under Scenarios 2, 4a, 4b, 4c, 5a, 5b, 5c (school closure scenarios), the homebound peak for adults was highest under Scenarios 5a, 5b, 5c, due to shelter-in-place, ranging from 5,377,886 to 5,379,960, followed by homebound peak of 584,235 under Scenario 2. Adults absent from work followed a similar pattern as homebound adults across all scenarios (*Supplementary Material Figure B1*).

**Figure 2:**
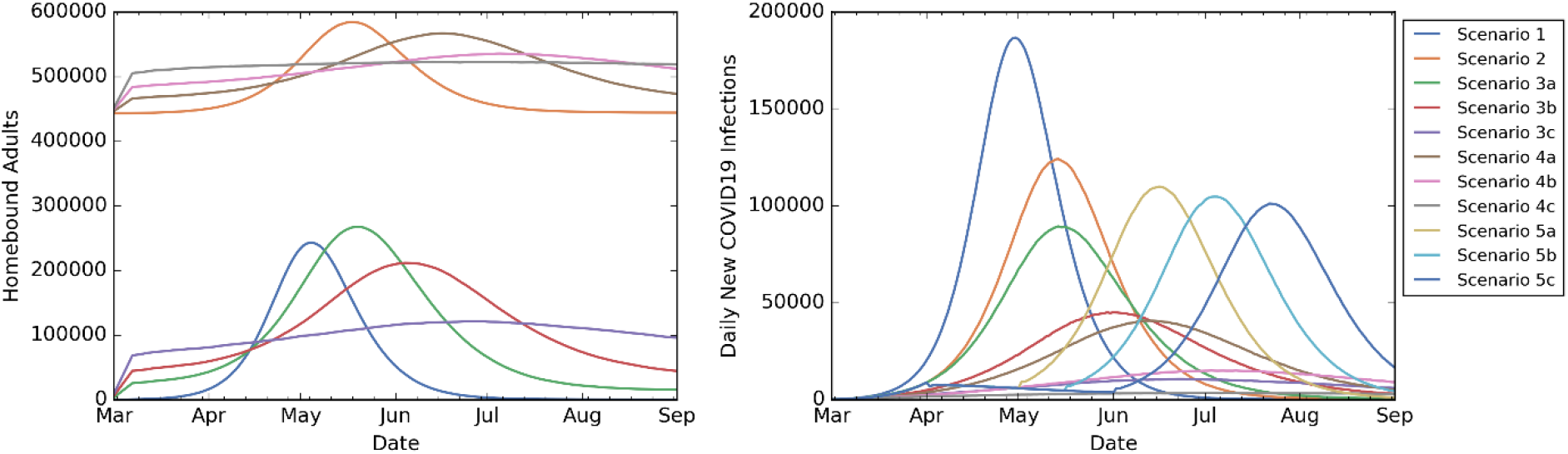
Homebound adults and daily new infections over time. Scenarios 2, 4a, 4b, 4c include school closure.

Higher compliance with voluntary quarantine reduced homebound peak for adults to 566,973, 535,559, 522,775 under Scenarios 4a, 4b, 4c, respectively (*Figure 2*), decreased the peak infection (in Scenarios 3a, 3b, 3c, 4a, 4b, 4c) by at least half, and delayed the peak day by 14-61 days compared to Scenario 1 (*Supplementary Material Table B2*).

*Figure 3* presents a comparison of the percentage of the population infected or dead and the percentage of days adults homebound. The percentage of the population infected was 60.09% under Scenario 1 (no intervention) and 51.69% under Scenario 2 (school closure only). The percentage of the population infected reduced to a range of 11.68-44.15% under Scenarios 3a, 3b, 3c (voluntary quarantine) and 4.53-31.07% under Scenarios 4a, 4b, 4c (voluntary quarantine with school closure). The percentage of days adults homebound was 0.72% under Scenario 1 and 7.16% under Scenario 2 (school closure only). The percentage of days adults homebound ranged from 1.36-1.63% and 7.62-7.74% under Scenarios 3a, 3b, 3c (voluntary quarantine) and Scenarios 4a, 4b, 4c (voluntary quarantine with school closure), respectively. Compared to Scenario 2 (school closure only), Scenarios 5a, 5b, 5c (shelter-in-place with school closure) reduced the percentage of the total population infected from 51.69% to 48.11-50.55% but more than doubled the percentage of days adults homebound to a range of 18.92-30.66%.

**Figure 3:**
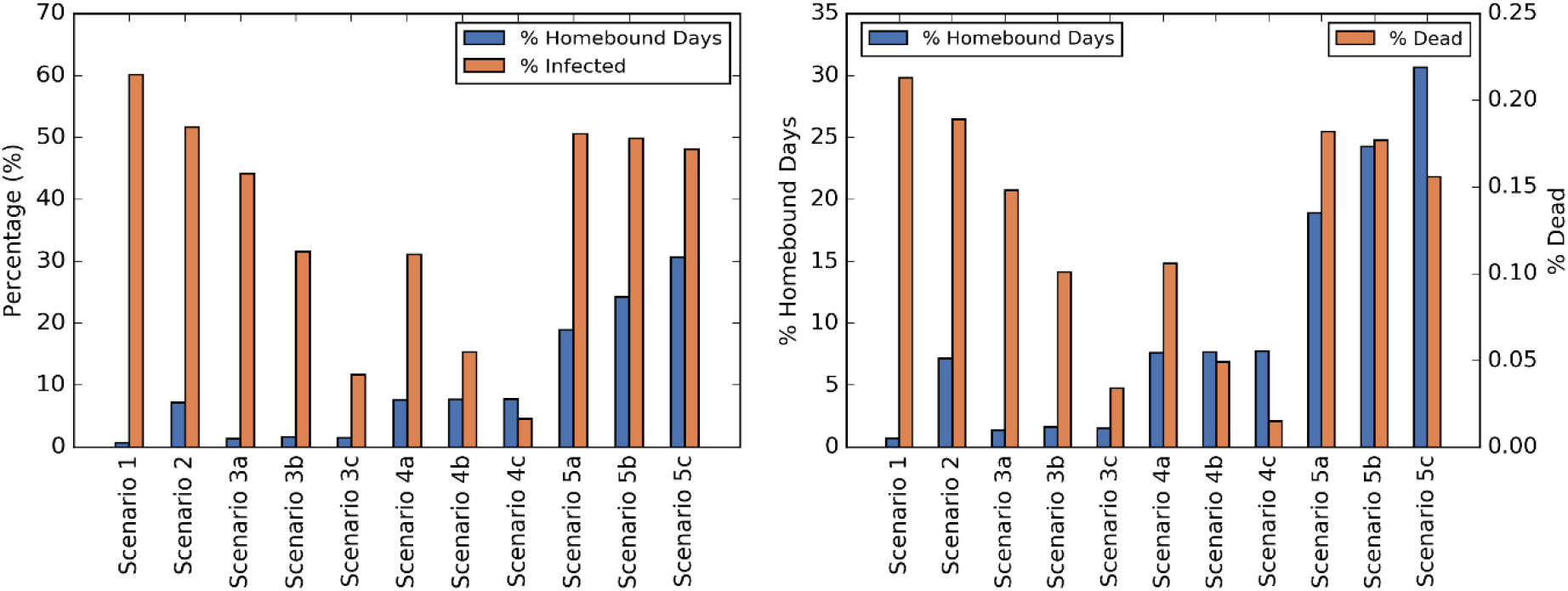
Percentage of days adults homebound compared to the percentage of the population infected (left figure) and dead (right figure).

*Supplementary Material Table B3* provides the percentage of days children, youth, adults, and elderly are homebound across all scenarios.

*Figure 4* presents the homebound days to prevent an infection or death. The homebound days to prevent an infection was 87 under Scenario 2 (school closure only) and over 200 under Scenarios 5a, 5b, 5c (shelter-in-place with school closure). The homebound days to prevent an infection was 1.9, 3.7, 4.7 under Scenarios 3c, 3b, 3a (voluntary quarantine), respectively, versus 14.4, 17.8, 27.3 under Scenarios 4c, 4b, 4a (voluntary quarantine with school closure), respectively. The homebound days to prevent a death was 30,650 under Scenario 2 (school closure only) and over 60,420 under Scenarios 5a, 5b, 5c (shelter-in-place with school closure). The homebound days to prevent a death was 500, 928, 1,130 under Scenarios 3c, 3b, 3a (voluntary quarantine), respectively, versus 4,050, 4,840, 7,360 under Scenarios 4c, 4b, 4a (voluntary quarantine with school closure), respectively.

**Figure 4:**
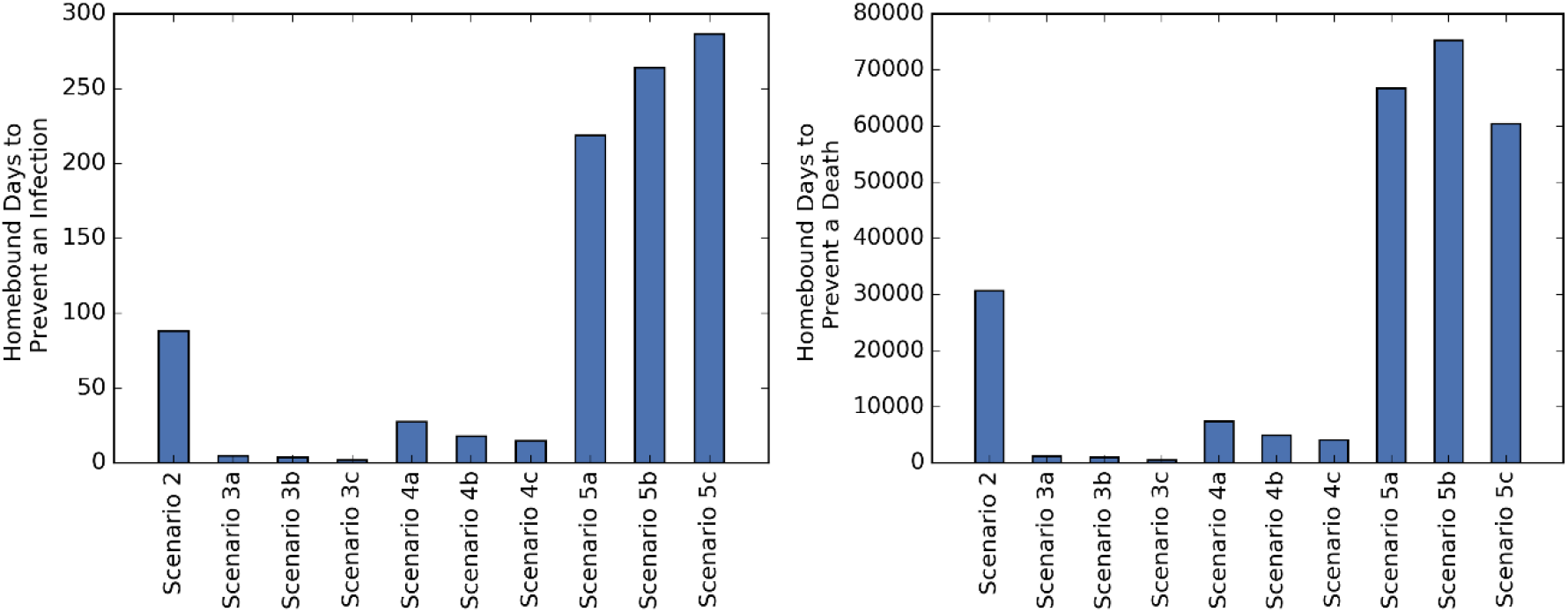
Homebound days to prevent an infection (left figure) or a death (right figure).

*Supplementary Material Table B4* presents the homebound and inactive peak percentages for children, youth, adults, elderly, and the total population. Increasing voluntary quarantine compliance, regardless of school closure, decreased the homebound and inactive peak percentage for adults, elderly, and the total population. *Supplementary Material Figures B2 and B3* present the homebound peak broken down by statuses for adults and elderly and for children and youth, respectively.

*Supplementary Material Figure B4* shows the percentage distribution of statuses (at home childcare, voluntary quarantine, symptoms) for the homebound peak for adults. At the homebound peak, among homebound adults: (i) Under Scenarios 2, 4a, 4b, 4c (school closure scenarios without shelter-in-place), 0.33%-27.26% and 72.75%-83.47% were symptomatic or providing at home childcare, respectively. (ii) Under Scenarios 3a, 3b, 3c (non-school closure scenarios), 3.61%-26.59% and 0.89%-4.04% were symptomatic or providing at home childcare, respectively. (iii) Under no intervention, 89.99% and 10.01% were symptomatic or providing at home childcare, respectively.

*Supplementary Material Tables B5-B7* summarize the impact of voluntary quarantine, school closure and shelter-in-place by comparing the percentage difference between a pair of scenarios in terms of the homebound days (for children, youth, adult and elderly populations), cumulative infections, and deaths.

## DISCUSSION

Many governments are faced with difficult decisions about when and how quickly to lift social distancing restrictions and reopen their economies; hence, it is crucial to analyze the benefits of NPIs in decreasing the spread of COVID19 versus the economic and social consequences considering the people who become homebound due to illness or due to complying with NPIs. In the state of Georgia, school closure began on March 16^th^ and a shelter-in-place order was enacted from April 3^rd^ to April 30^th^, and the reopening of the state continued gradually since then. The number of new COVID19 confirmed cases in Georgia have rapidly increased since early June.

This study focused on the public health benefits versus the need to refrain from societal or workplace interactions due to NPIs. The COVID19 pandemic led to widespread school closure and shelter-in-place orders in the United States^27,28^. Despite the potential public health benefits, there were many concerns about the economic impacts of shelter-in-place^10-13^ and the disruptive effects of school closures on the education of children and youth^14,21-23,29^. This study analyzed and compared several NPI scenarios, including combinations of school closure, voluntary quarantine, and shelter-in-place, with varying compliance levels and durations, as well as baseline scenarios of no intervention (Scenario 1) and school closure only (Scenario 2).

School closure alone had limited impact on reducing the spread of COVID19. Compared to no intervention, school closure only reduced the percentage of the population infected by less than 10% while more than doubling the peak number of adults homebound and causing nearly 450,000 work absences, the majority of which due to the need to provide at home childcare.

Shelter-in-place combined with school closure (Scenarios 5a-5c) temporarily slowed down the infection spread and delayed the peak, but had little impact on the magnitude of the peak and the cumulative number of infections and deaths, which were similar to that observed in the school closure only scenario. However, under Scenarios 5a-5c, the peak number of homebound adults was 9-44 times larger than all other intervention scenarios. Hence, the limited positive public health impact of shelter-in-place came at a very high societal cost.

Under voluntary quarantine (Scenarios 3a, 3b, 3c) the percentage of the population infected was 11.68%-44.15% (compared to 60.09% under no intervention), with the peak number of adults homebound being 267,566 under low, 211,695 under medium, and 121,346 under high compliance. Under voluntary quarantine combined with school closure (Scenarios 4a, 4b, 4c) the percentage of the population infected was 4.53%-31.06% (compared to 51.69% under school closure only), with the peak number of adults homebound being 566,973 under low, 535,559 under medium, and 522,775 under high compliance. Higher levels of voluntary quarantine compliance decreased the percentage of the population infected and the peak number of adults homebound (or absent from work).

Voluntary quarantine compliance provided the greatest benefits in terms of the reduction in infections and deaths compared to the number of adults homebound. Compared to school closure only, voluntary quarantine combined with school closure yielded up to a 92% decrease in cumulative infections and deaths while homebound days increased by at most 8% for adults, 6% for elderly and 1.5% for the total population. Under voluntary quarantine scenarios, the number of homebound days to prevent an infection or death was 3-154 times lower than that of all other scenarios.

## CONCLUSION

While large-scale interventions such as shelter-in-place temporarily slow down the infection spread, they are highly disruptive to the society and their public health impact is limited unless they are imposed for long durations of time, with high compliance levels, or followed by additional interventions.

Targeted interventions such as voluntary quarantine or voluntary quarantine combined with school closure significantly reduce the infection spread without causing a social and economic disruption as in the case of an extended shelter-in-place.

Strong public messaging should continue about voluntary quarantine, voluntary shelter-in-place (if possible), as well as other practices of physical distancing and the usage of facemasks.

Some of the conclusions of the study may be generalized to other states/countries that have geographic and population characteristics similar to the state of Georgia. The model and analysis would need to be adjusted for other pandemics; for example, COVID19 leads to fewer adverse health outcomes in younger populations and this may explain why school closure have a lesser impact on reducing infection spread.

### Limitations

If facemask usage was also considered in the NPI scenarios, the relative reduction in the number of cases and deaths could be higher compared to baseline scenarios. The simulation was populated with data from the state of Georgia and the results presented may not apply to other states or regions which have significantly different population characteristics or density.

## Data Availability

Publicly available data.

https://dph.georgia.gov/covid-19-daily-status-report

## Acknowledgements

The authors of this paper are thankful to state representatives for sharing multiple data sources from the cases confirmed in Georgia to date. The authors are also thankful to Melody Shellman, Hannah Lin, Ethan Channel, Pravara Harati, April Yu Zhuoting, Gabriel Siewert, and Christopher Stone for supporting various parts of the projects.

